# Development and Validation of a Deep Learning Model for Automated View Classification of Pediatric Focused Assessment with Sonography for Trauma (FAST)

**DOI:** 10.1101/2020.10.14.20206607

**Authors:** Aaron E. Kornblith, Newton Addo, Ruolei Dong, Robert Rogers, Jacqueline Grupp-Phelan, Atul Butte, Pavan Gupta, Rachael A Callcut, Rima Arnaout

## Abstract

The pediatric Focused Assessment with Sonography for Trauma (FAST) is a sequence of ultrasound views rapidly performed by the clinician to diagnose hemorrhage. One limitation of FAST is inconsistent acquisition of required views. We sought to develop a deep learning model and classify FAST views using a heterogeneous dataset of pediatric FAST. This study of diagnostic test developed and tested a deep learning model for view classification of archived real-world pediatric FAST studies collected from two pediatric emergency departments. FAST frames were randomly distributed to training, validation, and test datasets in a 70:20:10 ratio; each patient was represented in only one dataset to maintain sample independence. The outcome was the prediction accuracy of the model in classifying FAST frames and video clips. FAST studies performed by 30 different clinicians from 699 injured children included 4,925 videos representing 1,062,612 frames from children who were a median of 9 years old. On test dataset, the overall view classification accuracy for the model was 93.4% (95% CI: 93.3-93.6) for frames and 97.8% (95% CI: 96.0-99.0) for video clips. Frames were correctly classified with an accuracy of 96.0% (95% CI: 95.9-96.1) for cardiac, 99.8% (95% CI: 99.8-99.8) for thoracic, 95.2% (95% CI: 95.0-95.3) for abdominal upper quadrants, and 95.9% (95% CI: 95.8-96.0) for suprapubic. A deep learning model can be developed to accurately classify pediatric FAST views. Accurate view classification is the important first step to support developing a consistent and accurate multi-stage deep learning model for pediatric FAST interpretation.

## INTRODUCTION

Focused Assessment with Sonography for Trauma (FAST) is a point-of-care ultrasound diagnostic study, in which a sequence of thoracic and abdominal ultrasonographic views performed by the treating provider is used to rapidly identify free fluid (hemorrhage) secondary to blunt abdominal trauma. The FAST is beneficial because it avoids the use of ionizing radiation, decreases computed tomography (CT) usage, and improves outcomes in injured adults.(1) However, FAST has not been shown to improve outcomes in injured children, mainly because of variable diagnostic test characteristics.(2) The varying diagnostic test characteristics of FAST in children arise from patient characteristics, but also inconsistent provider experience in technical reliability in image acquisition and interpretation, leading to variable diagnostic accuracy and provider confidence.(3-5) Expertise in FAST improves the overall accuracy of the study, and therefore, the utility in potentially reducing unnecessary CT use.(6) However, barriers in education, training, skill maintenance, and quality assurance have perpetuated a lack of global FAST expertise.(3, 7, 8) This lack of expertise is often cited as the primary reason for FAST not being accurately implemented in clinical practice.(9-11) Therefore, it would be advantageous to develop novel methods of disseminating widespread expertise in pediatric FAST, as a potentially life-saving and radiation-sparing diagnostic tool for use in the treatment of injured children.

Deep learning has been shown to be useful in many aspects of clinical image processing including automatic classification of diagnostic ultrasound, but not yet in pediatric FAST.(12) View classification models are often the first step in creating robust and interpretable deep learning models for successful diagnostic ultrasound applications.(13, 14) View classification allows researchers insight into the feasibility of a deep learning approach, while also contributing to the understanding of a multistage model.(15, 16) Previous diagnostic ultrasound deep learning classification models, including those for echocardiography and radiology-performed diagnostic ultrasound, have utilized imaging data acquired from operators often with advanced training and accreditation in image acquisition.(13, 14, 17) Accredited operators are apt to derive standard sequences and higher quality image capture.(18, 19) In contrast, pediatric FAST represents a distinct challenge because image acquisition and interpretation are performed by treating providers with varying ultrasound training and skill levels. As such, provider-performed FAST studies have variability in image acquisition,(20) creating a layer of complexity when building a classification model. Therefore, we sought to evaluate the ability of a deep learning model to recognize point-of-care ultrasound features for view classification, in an effort toward eliminating barriers to clinical implementation of FAST in the treatment of injured children.

## MATERIALS AND METHODS

### Study Design & Setting

We collected a convenience sample of archived pediatric FAST studies performed on children during routine emergency department (ED) care at two institutions, UCSF Benioff Children’s Hospital Oakland (BCHO) and UCSF Benioff Children’s Hospital San Francisco (BCHSF). UCSF Benioff Children’s Oakland is a regional trauma referral center and pediatric emergency department with approximately 45,000 visits annually, and BCHSF, is a tertiary referral center and pediatric emergency department receiving approximately 18,000 visits annually. Each hospital’s point-of-care ultrasound quality assurance program stores and catalogs FAST studies. FAST studies from March 1, 2018, through January 1, 2020, were retrieved in raw Digital Imaging and Communications in Medicine (DICOM) format from each hospital’s Picture Archiving and Communication Systems (PACS). This project received review approval by the Human Research Protection Program Institutional Review Board (IRB) from the University of California, San Francisco and the UCSF Benioff Child’s Hospital Oakland. All data was held in a secure, high-performance computing environment in compliance with the IRB. Before processing, all images were deidentified according to the Health Insurance Portability and Accountability Act (HIPAA) Safe Harbor standards using a proprietary process developed at UCSF. Additionally, relevant clinical and trauma registry databases were queried from each site to be linked in a deidentified manner to the respective FAST study. All FAST studies were acquired with either the Sonosite Edge II (Fujifilm, Inc., Bothell, Washington) or the Sonosite X-Porte (Fujifilm, Inc., Bothell, Washington) with transducer frequencies between 13 to 1 Mhz.

### Subjects

We included FAST studies from patients younger than 18 years of age who were evaluated for a traumatic mechanism. FAST studies were excluded if FAST study and patient data could not be linked.

### Imaging Dataset and Preprocessing

An imaging dataset of the pediatric FAST studies from children with blunt abdominal trauma was created. A FAST study was defined as one or more video clips from a single patient. Each video clip represents one of the possible FAST views (Figure 1). FAST views include standard abdominal and thoracic sonographic views, including the cardiac views, thoracic views, abdominal upper quadrant views, and suprapubic views. The ultrasound systems capture still frames at a rate of 30 still frames per second. Video clips for review were then generated at a rate of 60 still frames per second from labeling. These FAST still frames were all then resized to squared portable network graphics (PNG) of 299 × 299 pixels and normalized to appear with a standardized intensity.

**FIGURE 1.**
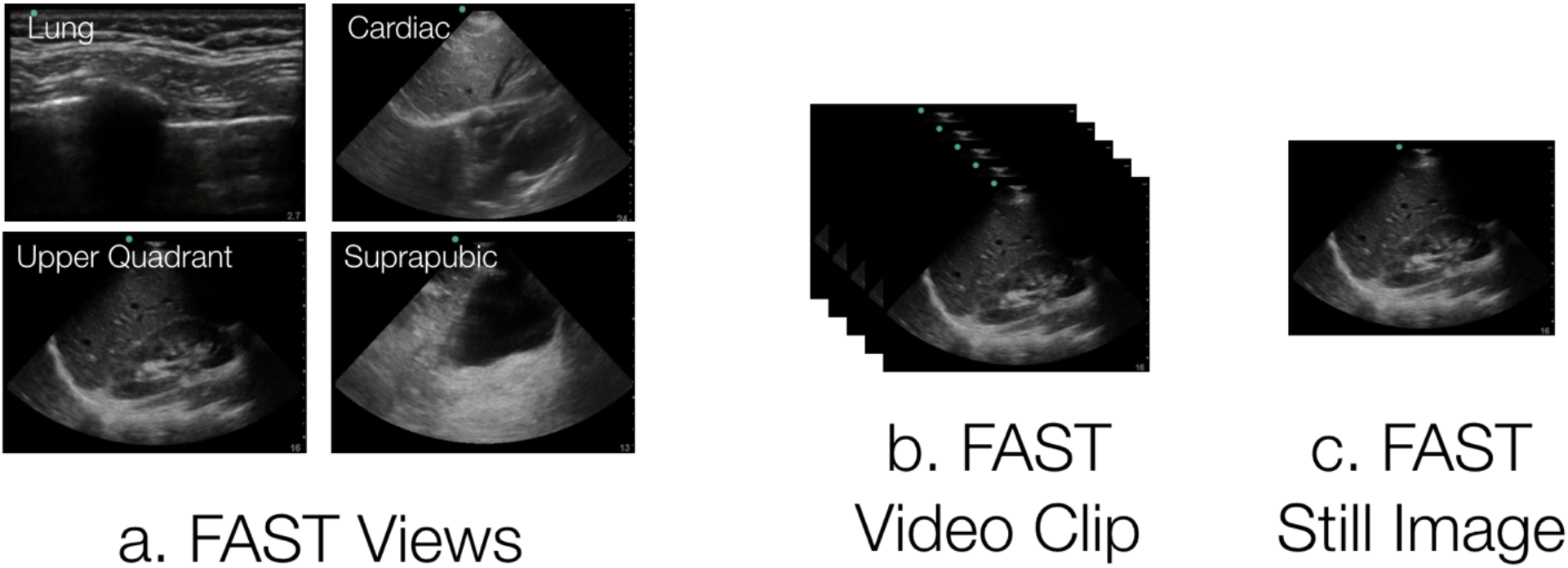
A FAST study is made up of a sequence of (a) FAST Views. Each FAST View is archived as a (b) FAST Video Clip(s). Each FAST Video Clip is made up of multiple (c) FAST Still frames.

### Image Video Labeling

All FAST studies were labeled by a fellowship-trained expert in pediatric FAST. FAST video clips were blindly labeled once as either: cardiac, thoracic, abdominal upper quadrant, or suprapubic. Where possible, the abdominal upper quadrant views also received a secondary label for laterality (corresponding to left [LUQ] or right [RUQ]) abdominal upper quadrants for a secondary sensitivity analysis). After labeling, FAST studies were randomly partitioned (Ratio: 70-20-10) into training, validation, and test datasets.(21) A training dataset refers to a sample of data used to fit the deep learning model. The validation dataset refers to a new set of data used to provide an unbiased evaluation of a model fit on the training dataset. The test dataset refers to a separate set of data used to provide an assessment of a final model fit. FAST video clips from a single patient could be included in only one dataset to maintain sample independence. A second ultrasound expert independently reviewed the test dataset labels to ensure reliability. The second expert was blinded to the results of the first expert.

### Model Architecture and Training

The deep learning model, ResNet-152, is a convolutional neural network (CNN) pre-trained on Imagenet, that reduces the time to computationally converge on a successful model and maximize the available computed feature set across a deep neural network. ResNet-152 is based on the extraction of defining features from objects within images. A one-cycle training was implemented with a batch size of 40 images and a final dropout of p=0.5. We utilized a predefined training technique including 12 epochs (complete passes over the dataset), with 8 epochs focused on the top layer of ResNet-152, and 4 epochs allowing changes to weights across the entire network.(22) Model optimization was implemented using an adaptive gradient descent method with weight decay and a flattened cross-entropy loss function.

### Data Augmentation

Spatial transformations and other standard image manipulations were incorporated in an additional candidate model to introduce artificial noise in the dataset to reduce the chance of model overfitting. The transformation techniques included: 1) rotation (0.75 probability of rotation 0 to 180 degrees right or left), 2) cutout (0.75 probability loss of up to 4 regions 10 to 160 pixels in size), 3) crop and resize (0.75 probability of a 299 × 299 pixel region selected from image scaled 0.75x to 2x original), and 4) adjustment of brightness and contrast (0.75 probability of a .05x to 0.95x brightness and/or 0.1x to 10x contrast change) to modify a subset of our images. This is a standard approach in computer vision tasks (15, 16) which we applied here to assess the classifier’s ability to handle increasingly varied images that differ significantly from the standard ultrasound presentation.

### Outcome Measures

The primary outcome measure was the deep learning model’s classification of FAST view by (1) video clip and (2) still frame on the independent test dataset. The secondary outcome tested the model’s accuracy for view classification with the addition of differentiation of the two abdominal upper quadrant views, left and right, by video clips and still frame. The reference standard was expert FAST interpretation. The time required for the model to make a prediction was also reported by study.

### Evaluation of Model Performance

Descriptive statistics were reported as frequencies and proportions to describe the sample study population and expert classification. In the portion of the studies reserved for testing model performance, the output labels for both single still frames and full video clips were compared to the expert’s manual label. For each still frame the model assesses, it independently outputs a “probability” of class membership for each of the four classes. The class with the highest probability score is assigned as the output label for a single still frame, and the video clips are assigned an output label by using a plurality classification from all the still frames generated from a single video clip. (13)

We calculated a confusion matrix and computed diagnostic accuracy measures for the model’s predicted classes. Model training and statistical analyses were performed using Python version 3.7 (Python Software Foundation) software, and R, version 3.6 (R Foundation for Statistical Computing). Class-specific accuracies are computed as the total proportion of true positive and true negatives for each predicted output class. Overall model accuracies are calculated as the total proportion of all correct classifications across all included views. F-scores were additionally computed as twice the harmonic mean of the positive predictive value (PPV) and sensitivity. Clopper-Pearson confidence intervals were also included for each proportion estimated. We sought to plot a visual representation of the transformed images as interpreted by the model. In a random sample of still images from the test dataset, we selected the 4096-dimensional vector output from the last fully connected network layer (FC7) before the model’s output prediction image. These high-dimension vectors were initially reduced to 50 dimensions with principal components analyses and then further to two dimensions with T-Distributed Stochastic Neighbor Embedding (t-SNE) to visualize dimensionality reduction.(23) Similarity among the model’s interpretation of the images is represented by the lower Euclidian distance between the points when plotted.(23, 24) We utilized gradient-weighted class activation mapping (Grad-CAM), a technique for the visual representation of the contribution of regions within an image to the model’s prediction.(25) The code used to generate this model and statistical analysis will be available upon request. However, ultrasound images are unavailable because of institutional regulation.

## RESULTS

We deidentified 701 FAST studies with recorded video clips. Two studies were excluded for missing linked patient-level data. 699 FAST studies were included in our analysis. There were a total of 4,925 video clips representing 1,062,612 still frames. The video clips were comprised of abdominal upper quadrant (45.8%), suprapubic (24.8%), cardiac (18%), and thoracic (12%). The majority of the FAST studies came from children less than 11 years of age, and one-third of the total studies came from children less than 6 years of age (Table 1). Sixty percent of the cohort were males. The majority of FASTs were obtained at the level 1 pediatric trauma center and were performed by 30 attending providers. There was no single attending provider that performed more than 11% of the FAST studies.

**TABLE 1.**
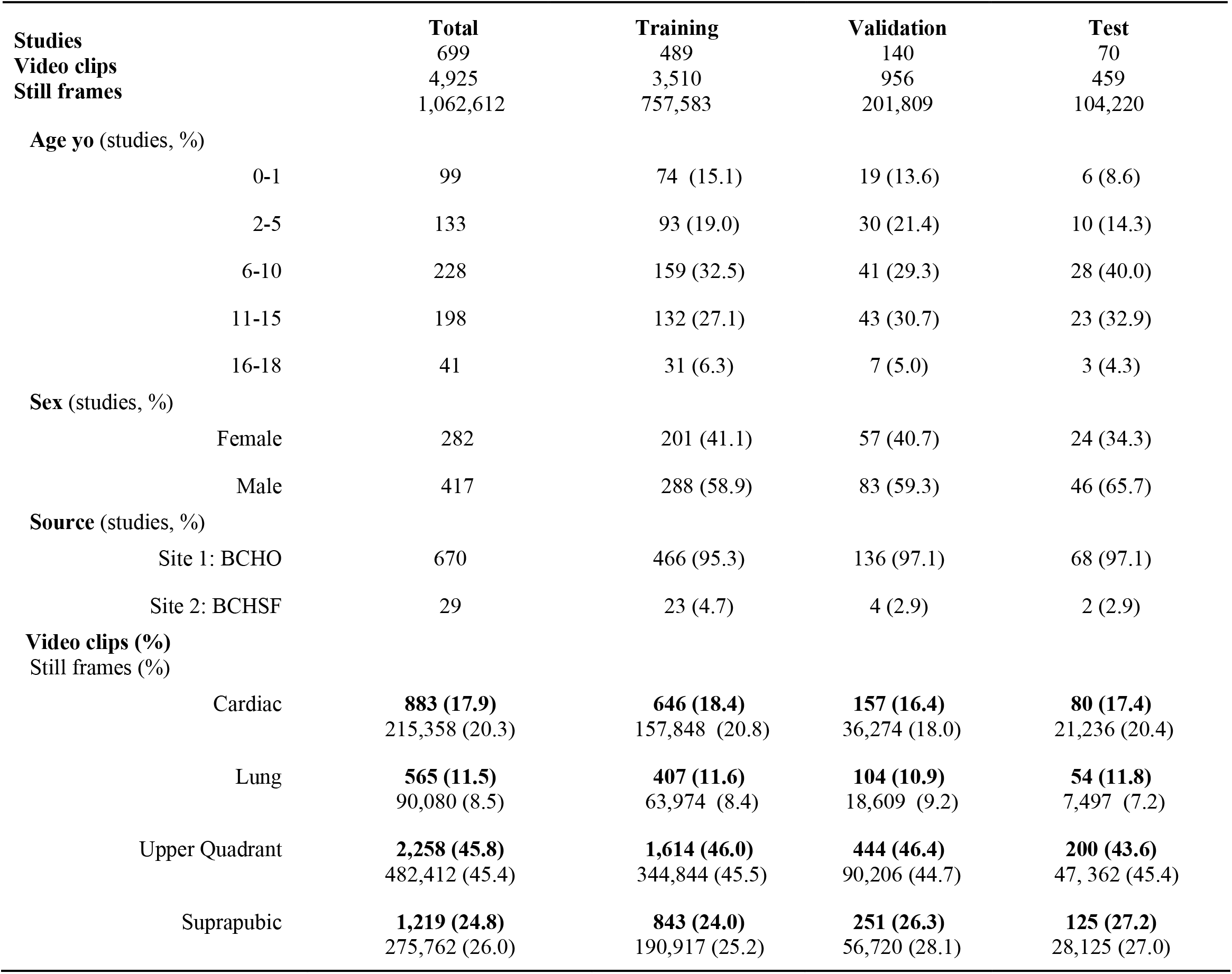
Comparison of study sample characteristics for development and testing of the pediatric FAST deep learning model.

The 699 FAST studies were divided into the training (70%), validation (20%), and test (10%) datasets (Table 1). Age, gender, source, and expert labeling were evenly distributed between the different datasets. The test dataset was made up of 70 FAST studies, 459 video clips, and 104,220 still frames, which had a slightly higher proportion of studies from children 6-10 years of age, females, and suprapubic video clips/still frames.

### FAST View Classifier

After training on 757,583 frames, we applied the model to the test dataset. The standard clock time needed to predict the view in 1000 still frames in this set was approximately 40 seconds. The overall model classification accuracy was 97.8% (96.0 - 99.0%) for video clips, and 93.4% (93.3 – 93.6%) for still frames (Table 2). The F-scores were 0.98 and 0.94 for video clips and frames, respectively. The model had view classification accuracies of video clips of 98.7%, 99.8%, 98.3%, and 98.9% for the cardiac, thoracic, abdominal upper quadrant, and suprapubic views, respectively. The model had view classification accuracies of still frames of 96.0%, 99.8%, 95.2%, 95.9% for the cardiac, thoracic, abdominal upper quadrant, and suprapubic views, respectively. The clustering analyses showed that the model could sort FAST still frames into groups according to FAST view (Figure 2). The model performed with similar accuracy on the augmented test dataset (Table 3). Based on two independent study experts, Grad-CAM indicated that the model received prediction contributions from relevant view classification sites (Figure 3).

**TABLE 2.**
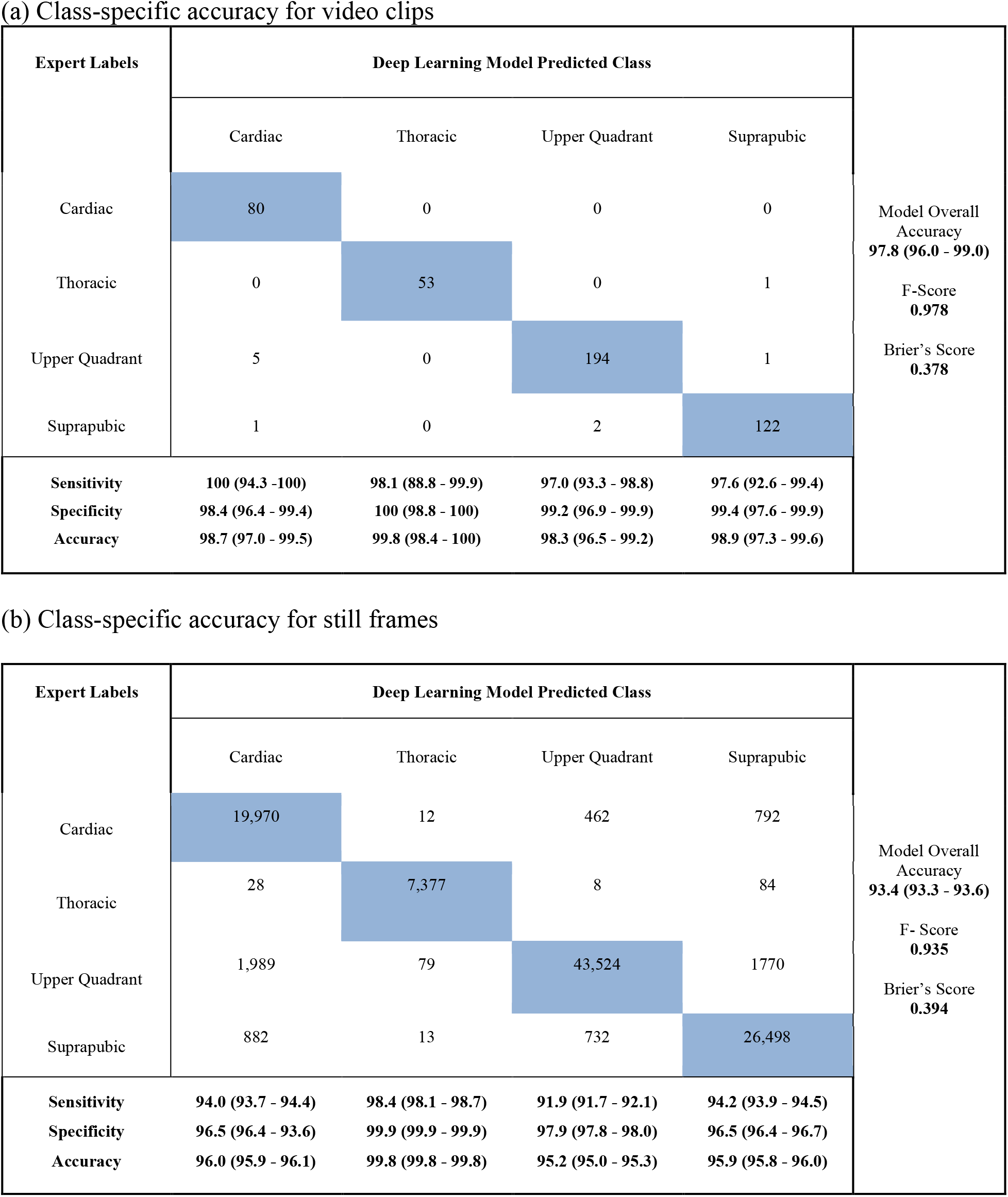
Pediatric FAST view classification accuracy by the deep learning model for (a) video clips and (b) still frames.

**(a) Class-specific accuracy for video clips**
| Expert Labels | Deep Learning Model Predicted Class |  |  |  |  |
| --- | --- | --- | --- | --- | --- |
|  | Cardiac | Thoracic | Upper Quadrant | Suprapubic |  |
| Cardiac | 80 | 0 | 0 | 0 | Model Overall Accuracy<br>**97.8 (96.0 - 99.0)**<br>F-Score<br>**0.978**<br>Brier's Score<br>**0.378** |
| Thoracic | 0 | 53 | 0 | 1 |  |
| Upper Quadrant | 5 | 0 | 194 | 1 |  |
| Suprapubic | 1 | 0 | 2 | 122 |  |
| Sensitivity | 100 (94.3 - 100) | 98.1 (88.8 - 99.9) | 97.0 (93.3 - 98.8) | 97.6 (92.6 - 99.4) |  |
| Specificity | 98.4 (96.4 - 99.4) | 100 (98.8 - 100) | 99.2 (96.9 - 99.9) | 99.4 (97.6 - 99.9) |  |
| Accuracy | 98.7 (97.0 - 99.5) | 99.8 (98.4 - 100) | 98.3 (96.5 - 99.2) | 98.9 (97.3 - 99.6) |  |

**(b) Class-specific accuracy for still frames**
| Expert Labels | Deep Learning Model Predicted Class |  |  |  |  |
| --- | --- | --- | --- | --- | --- |
|  | Cardiac | Thoracic | Upper Quadrant | Suprapubic |  |
| Cardiac | 19,970 | 12 | 462 | 792 | Model Overall Accuracy<br>**93.4 (93.3 - 93.6)**<br>F- Score<br>**0.935**<br>Brier's Score<br>**0.394** |
| Thoracic | 28 | 7,377 | 8 | 84 |  |
| Upper Quadrant | 1,989 | 79 | 43,524 | 1770 |  |
| Suprapubic | 882 | 13 | 732 | 26,498 |  |
| Sensitivity | 94.0 (93.7 - 94.4) | 98.4 (98.1 - 98.7) | 91.9 (91.7 - 92.1) | 94.2 (93.9 - 94.5) |  |
| Specificity | 96.5 (96.4 - 93.6) | 99.9 (99.9 - 99.9) | 97.9 (97.8 - 98.0) | 96.5 (96.4 - 96.7) |  |
| Accuracy | 96.0 (95.9 - 96.1) | 99.8 (99.8 - 99.8) | 95.2 (95.0 - 95.3) | 95.9 (95.8 - 96.0) |  |

**TABLE 3.**
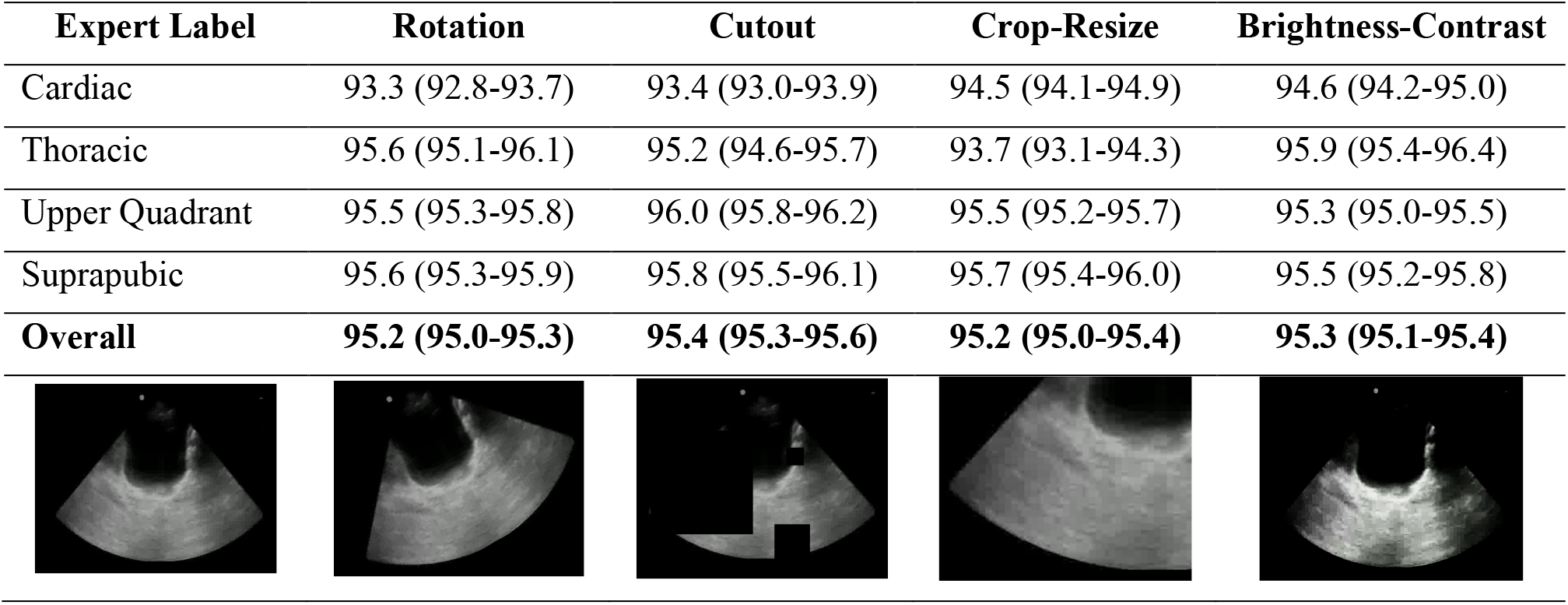
Pediatric FAST view classification accuracy (95% confidence interval) by the deep learning model for still frames after augmentation techniques.

| Expert Label | Rotation | Cutout | Crop-Resize | Brightness-Contrast |
| --- | --- | --- | --- | --- |
| Cardiac | 93.3 (92.8-93.7) | 93.4 (93.0-93.9) | 94.5 (94.1-94.9) | 94.6 (94.2-95.0) |
| Thoracic | 95.6 (95.1-96.1) | 95.2 (94.6-95.7) | 93.7 (93.1-94.3) | 95.9 (95.4-96.4) |
| Upper Quadrant | 95.5 (95.3-95.8) | 96.0 (95.8-96.2) | 95.5 (95.2-95.7) | 95.3 (95.0-95.5) |
| Suprapubic | 95.6 (95.3-95.9) | 95.8 (95.5-96.1) | 95.7 (95.4-96.0) | 95.5 (95.2-95.8) |
| **Overall** | **95.2 (95.0-95.3)** | **95.4 (95.3-95.6)** | **95.2 (95.0-95.4)** | **95.3 (95.1-95.4)** |

**FIGURE 2.**
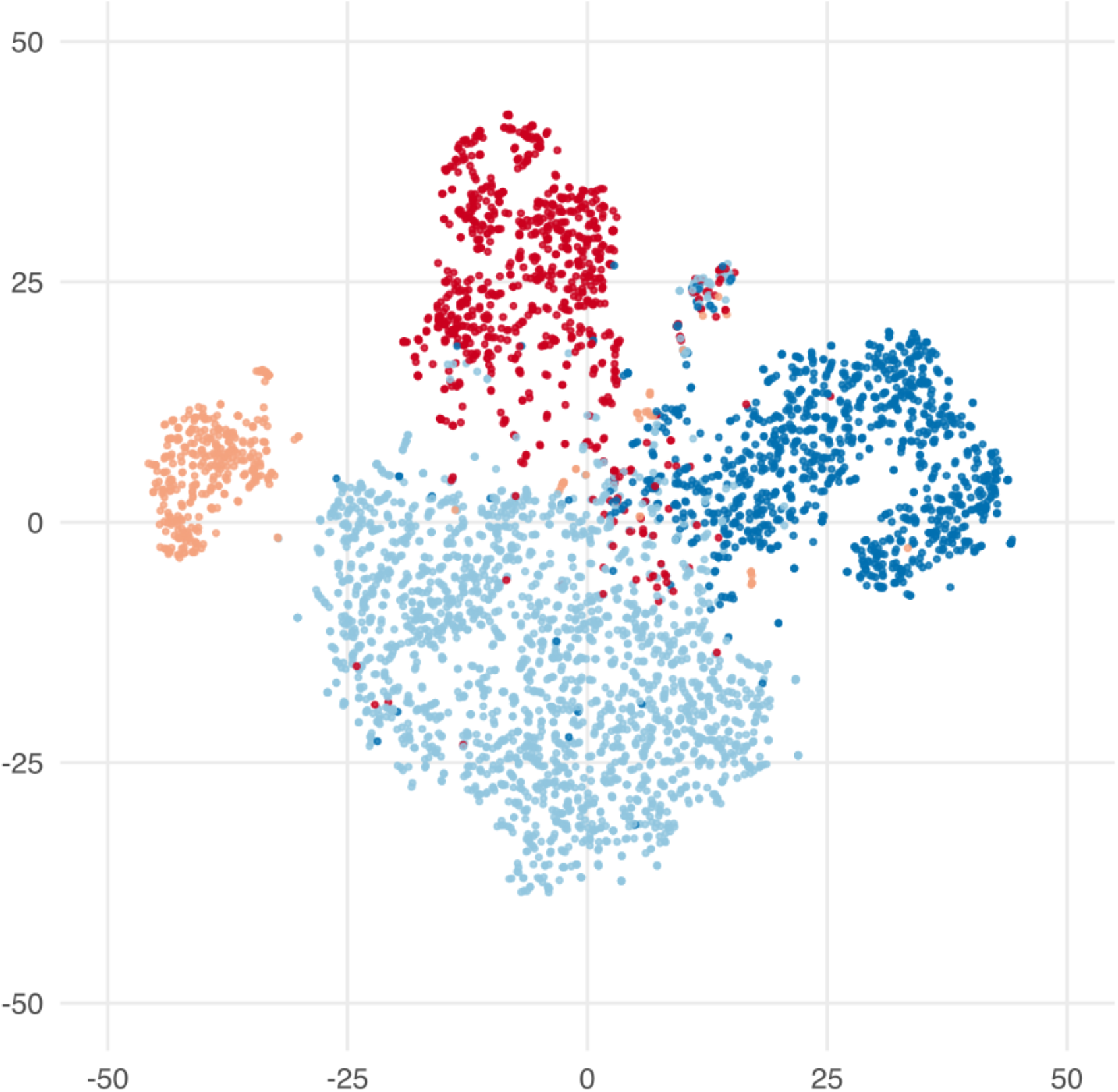
T-distributed stochastic neighbor embedding (t-SNE) plot. Clustering analysis of pediatric FAST views: Cardiac views (red), Thoracic views (peach), Abdominal upper quadrant views (light blue), suprapubic views (dark blue).

**FIGURE 3.**
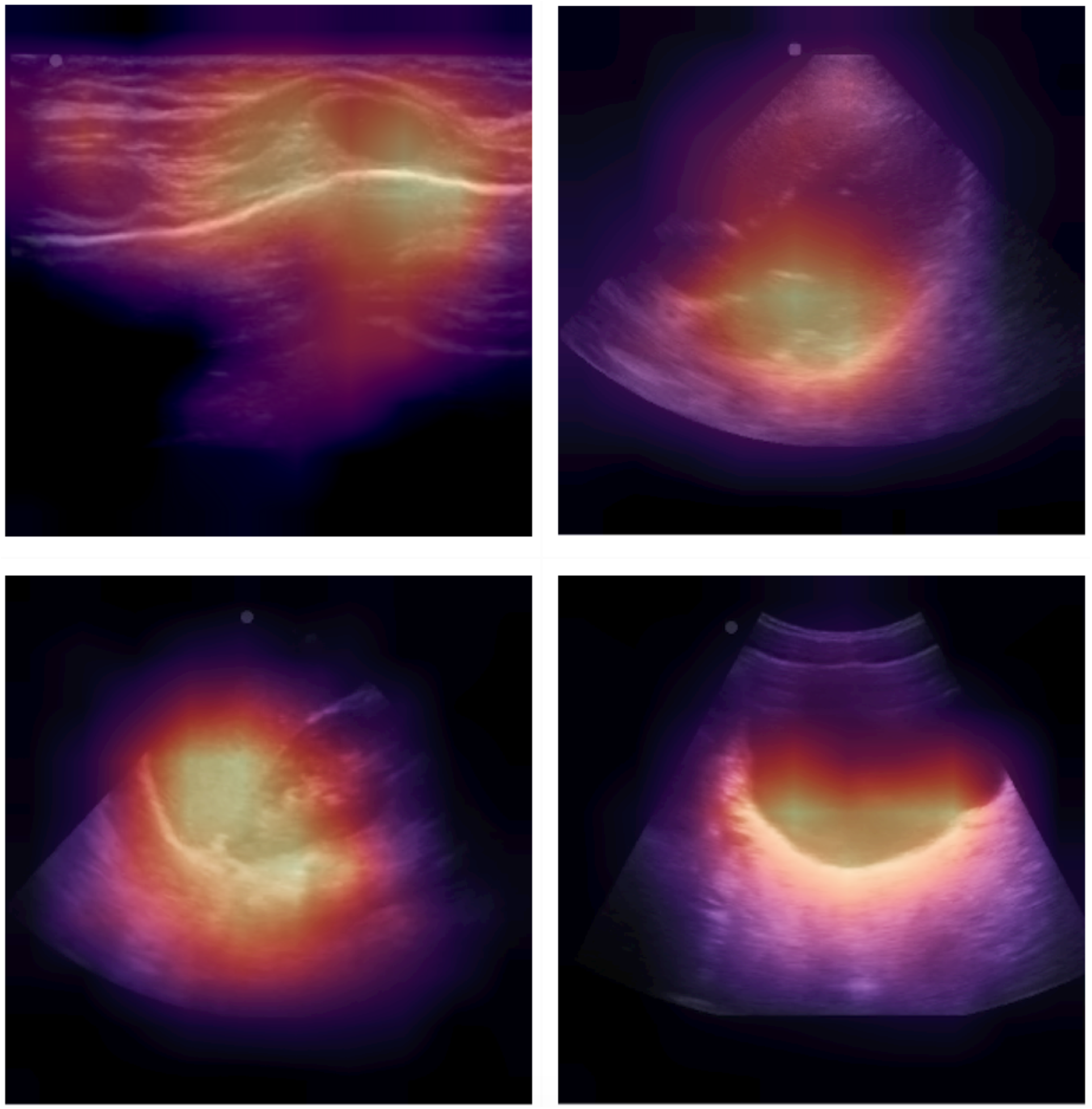
Gradient Class Activation Mapping (Grad-CAM). A. Thoracic view with highest contribution from costal-pleural interface. B. Cardiac view with highest contribution from left atrium. C. Upper Quadrant view with highest contribution from spleen-renal-diaphragm interface. D. Suprapubic view with highest contribution from posterior bladder wall.

We also evaluated a model with the view classification accuracy with the additional discernment of the abdominal right and left upper quadrant views. Forty-nine (24%) video clips of the upper abdominal views received a secondary expert label for right or left upper quadrant. 151 (76%) were given a definitive label, of which 69 (46%) right upper quadrant and 82 (54%) left upper quadrant views. The overall model view classification accuracies were 88.5% (85.0 – 91.5%) for video clips and 83.5% (83.3 – 83.8%) for still frames (Supplement Table A). The F-scores were 0.88 and 0.83 for video clips and still frames, respectively. In this five-class model, view classification accuracy was lower in the abdominal upper quadrant views than the other anatomic views.

### Discordant Results

The experts had 5 (1.1%) discordant classifications between one another with an almost perfect inter-rater reliability of Fleiss Kappa of 0.98 (0.97-0.99), suggesting near-perfect agreement between expert classification. Furthermore, 8 video clips were discordant between the expert reference standard and the model predication classification (Supplement Figure A). Four of the video clips were concordant between expert reviewers, but discordant between the model’s prediction classification. These four discordant results were expert:model interpretation as cardiac:abdominal upper quadrant, abdominal upper quadrant:cardiac (2), and abdominal upper quadrant:suprapubic, respectively. The remaining four discordant video clips were discordant between the two experts. Three of these expert discordant video clips were concordant between expert 2 and the model prediction. The remaining video clip was discordant between both experts and the model prediction.

## DISCUSSION

We found that a deep learning model can be trained to accurately perform view classification from a diverse set of real-world clinician-performed pediatric FAST studies. The model’s high accuracy implies the ability to parse out identifying features within an ultrasound image. Feature identification is the essential first step toward developing an accurate and interpretable deep learning multistage model to automate pediatric FAST. Notably, the model is accurate in separating abdominal FAST views, including the suprapubic view, which is the most sensitive abdominal view for intraabdominal hemorrhage detection in pediatric patients.(26)

The inclusion of real-world clinician-performed FAST studies strengthens our model’s generalizability by avoiding an idealized dataset. Further, this study’s strength is that our pediatric FAST dataset is a relatively large expert-labeled set of pediatric FAST video clips and still frames from children with suspicion for abdominal hemorrhage. Similarly, data were curated from a large group of clinician-performed FAST studies from 30 providers with varying levels of FAST training and expertise. This dataset is also unique given the relatively young patient population, with greater than 30% of children less than six years of age.

Our model’s pediatric FAST view classification accuracy suggests that a deep learning model can identify pertinent features to achieve class predication accuracy nearly as well as an expert. There is no clear association of the number of FAST studies one needs to perform for expertise gained.(27) Furthermore, the FAST was accepted by the trauma community in the 1990s (28, 29), but barriers to training and implementation still exist, suggesting a need for alternative methods to develop its use.(20) Pediatric FAST is a technically complex study because of a child’s compact anatomy. Previous reports of FAST show excellent reliability for view classification between expert FAST providers, but only moderate agreement for less experienced users.(30) Our model performed with similar accuracy to that of an expert, suggesting deep learning may offer a possible solution for variability between providers with inconsistent expertise in FAST.

Deep learning classifiers have been trained and tested in different diagnostic ultrasound applications, including echocardiography view classification and lesion detection within the liver and breast.(12, 13, 15) However, FAST represents a complex problem because of the variability in the expertise of FAST operators and inconsistency in the quality of images obtained. In contrast to other deep learning classifiers, FAST is performed by the treating provider instead of a certified ultrasonography technician, and real-time clinical decisions are made from the results. Furthermore, these studies are also conducted at a fast pace within a high stakes environment that is prone to error.(31, 32) The inclusion of a heterogeneous group of FAST operators adds natural variation to the dataset and increases the complexity of the task.

Our findings build on previous work, demonstrating that machine learning models can identify pertinent features from FAST. Specifically, Sjogren et al. demonstrated feasibility in using basic machine learning techniques, including support-vector machine models, in interpreting adult FAST.(33) The model achieved 100% (95% CI, 69 −100%) sensitivity and 90% (95% CI, 56-100%) specificity for free fluid detection using only the abdominal right upper quadrant view from 10 positive and 10 negative adult FAST studies. However, this study utilized a labor-intensive annotation method, a basic machine learning approach, a small number of FAST studies, and may not be adaptable to pediatric FAST, where post-traumatic abdominal pathology displays wide variability. In contrast, our study demonstrates that less labor-intensive expert labeling, along with an advanced deep learning technique, can also perform with a high degree of accuracy. Our view classification deep learning model also suggests that these techniques are well-suited for additional development of a multistage free-fluid classifier to interpret the presence or absence of free fluid representing intraabdominal hemorrhage for the pediatric FAST (1, 34, 35).

One of this study’s goals was to develop automated processes and methods to reduce the need for human input. Human input in dataset curation is laborious, subject to human error, resource-intensive, and is a rate limiting step in high-quality, in-depth learning model development.(12) Our automated image preprocessing pipeline and view classification model can accurately expedite the development of larger and more robust datasets for the future multistage FAST models.(36) Our imaging preprocessing pipeline masked extraneous data outside the ultrasound viewing area to avoid training from non-clinical relevant features.

Additionally, our preprocessing pipeline included data augmentation techniques to simulate image variation outside the standard differences in image quality to avoid overfitting the model. Similar to our image preprocessing, we developed a system optimized for discordant frame evaluation in which experts could quickly review images (Supplement Figure A). This discordant image review process allows for a rapid review and understanding of model inaccuracy.

We recognize there are limitations to our study design. First, this study’s primary outcome was to develop an accurate deep learning model for view classification of pediatric FAST instead of pathology identification. However, recognizing relevant features within pediatric FAST still frames ensures our preprocessing pipeline is working and that the model distinguishes relevant features. Second, in our secondary analysis, the accuracy declined once the model was tasked to discriminate between right and left abdominal upper quadrant views. These two views are similar in sonographic appearance, which may be why the model has difficulty in classification, but could also represent the difficulty in the expert’s interpretation of right from left. Similar to previous studies of FAST view classification, the abdominal upper quadrant view includes both right and left because of the similarity of features and location of free fluid detection, which may be difficult for clinicians to define.(30)

Third, our model’s generalizability is limited by using a small group of FAST experts as the reference, and the majority of images came from a single institution and one ultrasound manufacturer. Previous studies have shown a strong agreement between experts for FAST study view classification.(30) Our team created a series of augmentations to simulate a more robust set of scenarios to account for homogenized institutional and manufacturer data. These simulations and reviewable frameworks will be necessary for future models and will allow our team to expand to include more extensive, more diverse populations. These augmentation methods will be important for future work as they require less effort without compromising accuracy.(37)

## CONCLUSION

In conclusion, we demonstrate that a deep learning model can be developed to provide an accurate view classification for a heterogeneous set of pediatric FAST video clips and still frames. This model suggests that our in-depth learning approach recognizes discerning imaging features within the pediatric FAST. Accurate view classification is the important first step to support developing a consistent and accurate multi-stage deep learning model for pediatric FAST interpretation.

## Data Availability

Availability of data and material: This patient-level data is protected under the University of California, San Francisco and will not be available for sharing.
Code availability: The code for this publication will be available to enable external researches to test model results against new data via institutional repository.

## ACKNOWLEDGEMENTS

The authorship team would like to acknowledge Drs. Nathan Teismann and Ashkon Shaahinfar for their support in data collection.

