## Supplement Table A for "Development and Validation of a Deep Learning Model for Automated View Classification of Pediatric Focused Assessment with Sonography for Trauma (FAST)"

**Supplemental TABLE A. Pediatric FAST view classification accuracy by the deep learning model for (a) video clips and (b) still frames.**

(a) Video clips (410)

| **Expert Labels** | **Deep Learning Model Predicted Class** | | | | | Model Overall Accuracy  **88.5 (85.0- 91.5)**  F-Score  **0.884**  Brier’s Score  **0.286** |
| --- | --- | --- | --- | --- | --- | --- |
|  | Cardiac | Thoracic | Left Upper Quadrant | Right Upper Quadrant | Suprapubic |  |
| Cardiac | 80 | 0 | 0 | 0 | 0 |  |
| Thoracic | 0 | 53 | 0 | 0 | 1 |  |
| Left Upper Quadrant | 3 | 0 | 64 | 14 | 1 |  |
| Right Upper Quadrant | 0 | 0 | 25 | 44 | 0 |  |
| Suprapubic | 1 | 0 | 2 | 0 | 122 |  |
| **Sensitivity**  **Specificity**  **Accuracy** | **100 (94.3 - 100)**  **98.8 (96.7 - 99.6)**  **99.0 (97.4 - 99.7)** | **98.1 (88.8 - 99.9)**  **100 (98.7 - 100)**  **99.8 (98.4 - 100)** | **78.0 (67.3 - 86.1)**  **91.8 (88.1 - 94.4)**  **89.0 (85.5 - 91.8)** | **63.8 (51.3 - 74.7)**  **95.9 (93.1 - 97.7)**  **90.5 (87.1 - 93.1)** | **97.6 (92.6 - 99.4)**  **99.3 (97.2 - 99.9)**  **98.8 (97.0 - 99.6)** |  |

(b) Still frames (92,498)

| **Expert Labels** | **Deep Learning Model Predicted Class** | | | | | Model Overall Accuracy  **83.5 (83.3 – 83.8)**  F-Score  **0.833**  Brier’s Score  **0.299** |
| --- | --- | --- | --- | --- | --- | --- |
|  | Cardiac | Thoracic | Left Upper Quadrant | Right Upper Quadrant | Suprapubic |  |
| Cardiac | 19,982 | 12 | 312 | 130 | 800 |  |
| Thoracic | 28 | 7,377 | 8 | 0 | 84 |  |
| Left Upper Quadrant | 1041 | 4 | 14,335 | 3,693 | 771 |  |
| Right Upper Quadrant | 210 | 0 | 6,125 | 9,032 | 429 |  |
| Suprapubic | 884 | 13 | 634 | 49 | 26,545 |  |
| **Sensitivity**  **Specificity**  **Accuracy** | **94.1 (93.8 - 94.4)**  **97.0 (96.8 - 97.1)**  **96.3 (96.2 - 96.4)** | **98.4 (98.1 - 98.7)**  **100 (100 - 100)**  **99.8 (99.8 - 99.9)** | **72.2 (71.6 - 72.9)**  **90.3 (90.0 - 90.5)**  **86.4 (86.2 - 86.6)** | **57.2 (56.4 - 57.9)**  **94.9 (94.8 - 95.1)**  **88.5 (88.3 - 88.7)** | **94.4 (94.1 - 94.6)**  **96.8 (96.6 - 96.9)**  **96.0 (95.9 - 96.2)** |  |
