## Supplement Figure A for "Development and Validation of a Deep Learning Model for Automated View Classification of Pediatric Focused Assessment with Sonography for Trauma (FAST)"

### **Supplemental FIGURE A.** Interpretable review for view class-accuracy for concordant and discordant predictions between expert label and deep model prediction with a confidence probability.

a. Concordant prediction

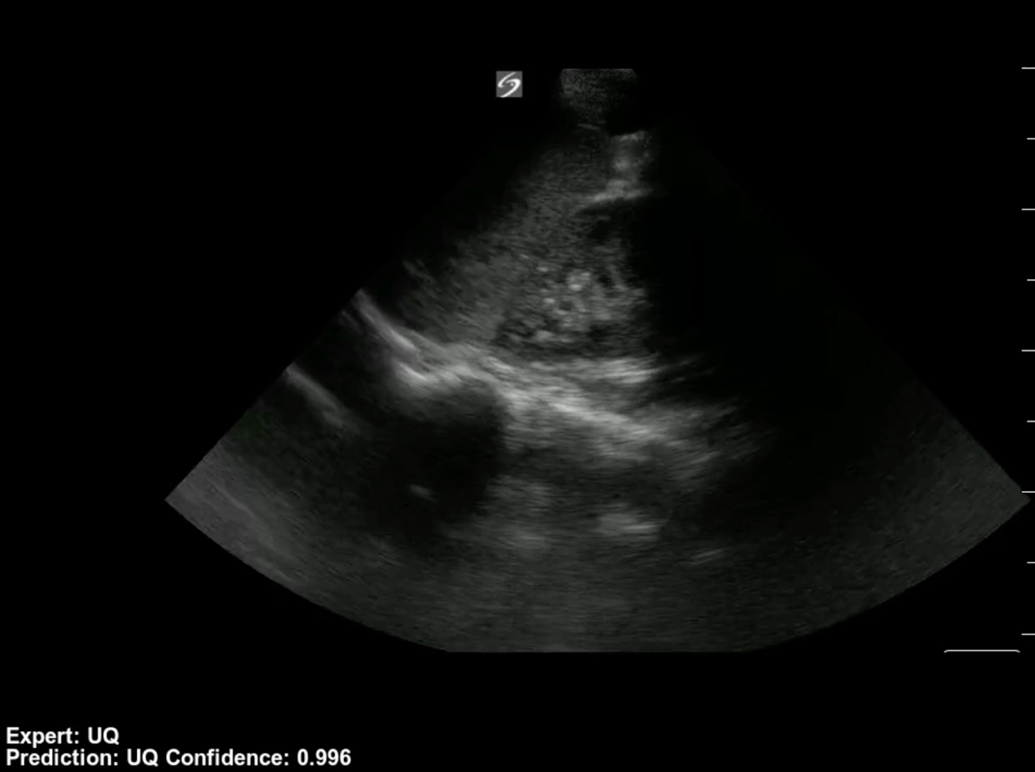

b. Discordant prediction because expert label was mislabeled.

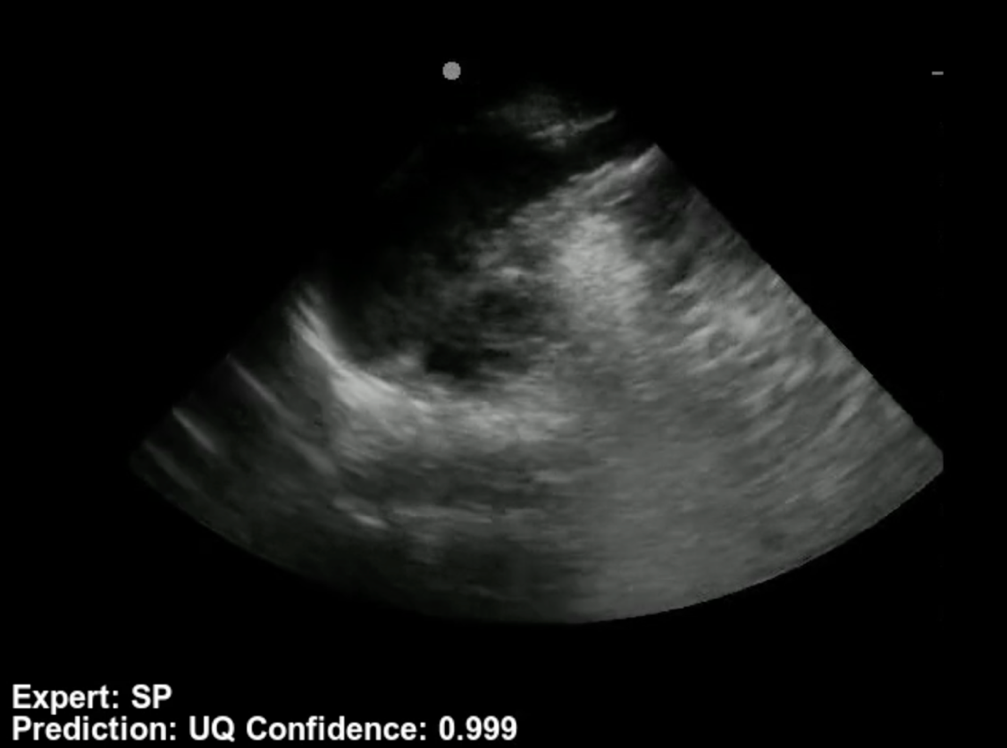

c. Discordant prediction because poor quality image. Note the prediction probability was low (0.36).

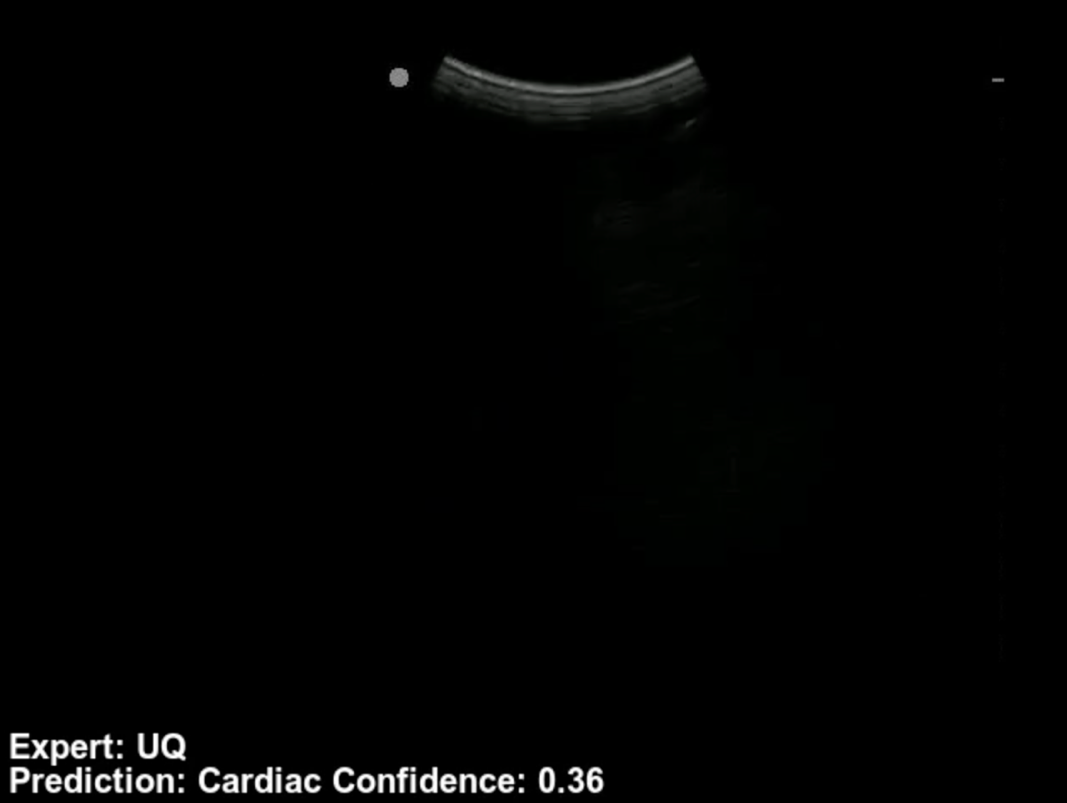
